# SARS-CoV-2 show no infectivity at later stages in a prolonged COVID-19 patient despite positivity in RNA testing

**DOI:** 10.1101/2021.03.18.21253228

**Authors:** Xiu-Feng Wan, Cynthia Y. Tang, Detlef Ritter, Yang Wang, Tao Li, Karen Segovia, Martina Kosikova, Marc Johnson, Hyung Joon Kwon, Hang Xie, Richard D. Hammer, Jane A. McElroy, Aws Hamid, Natalie D Collins, Jun Hang, Simone Camp

## Abstract

Inpatient COVID-19 cases present enormous costs to patients and health systems. Many hospitalized patients may still test COVID-19 positive, even after resolution of symptoms. Thus, a pressing concern for clinicians is the safety of discharging these asymptomatic patients if they have any remaining infectivity. This case report explores the viral viability in a patient with persistent COVID-19 over the course of a two-month hospitalization. Positive nasopharyngeal swab samples, analyzed by quantitative reverse transcription polymerase chain reactions (qRT-PCR), were collected and isolated in the laboratory, and infectious doses were analyzed throughout the hospitalization period. The patient experienced waning symptoms by hospital day 40 and had no viable virus growth in the laboratory by hospital day 41, suggesting no risk of infectivity, despite positive RT-PCR results, which prolonged his hospital stay. Notably, this case showed infectivity for at least 24 days from disease onset, which is longer than the discontinuation of transmission-based precautions recommendation by CDC. Thus, our findings suggest that the timeline for discontinuing transmission-based precautions may need to be extended for patients with prolonged illness. Additional large-scale studies are needed to draw definitive conclusions on the appropriate clinical management for these patients.

## INTRODUCTION

On March 11, 2020, the World Health Organization declared the coronavirus disease 2019 (COVID-19) a pandemic. COVID-19, caused by order *nidovirales*, family *coronaviridae*, genus *betacoronavirus*, species *severe acute respiratory syndrome coronavirus 2* (SARS-CoV-2), has cause more than 27 million laboratory-confirmed infections worldwide as of February 14, 2021^1^. Inpatient COVID-19 hospitalizations were projected to cost the US healthcare system up to $16.9 billion in 2020^2^ and imposes a large financial burden to individual patients^3^. The median hospital stay for COVID-19 is 10-13 days^4,5^. Many hospitalized patients with prolonged viral shedding may still test COVID-19 positive, even after resolution of symptoms, causing a prolonged hospitalization. Thus, a pressing concern for clinicians is the safety of discharging these asymptomatic patients if they have any remaining infectivity.

As of August 2020, the Center for Disease Control and Prevention (CDC) recommends the following guidelines for the discontinuation of transmission-based precautions for persons with severe or critical illness: patients up to 20 days after symptom onset, at least 24 hours after the last fever, and improved symptoms^6^. The CDC no longer recommends test-based strategies due to prolonged and detectable shedding in patients that no longer have infectivity^6^. In this case report, we present a patient with critical severity of COVID-19 disease who was still shedding infectious viruses at 24 days after symptom onset during his two-month long hospitalization.

## METHODS

### Ethics statement

This study was approved by the University of Missouri-Columbia Institutional Review Board (#2023844) and at the Institutional Biosafety Committee Biosafety Level 3 (#20-14).

### Sample Collection

The patient’s clinical observations were documented at least twice daily and multiple nasopharyngeal swabs and plasma samples were collected and tested to determine viral loads and Nab titers. Periodic national early warning scores (NEWS) were assessed. A score of seven or higher identifies high risk patients requiring activation of a medical emergency team. The patient’s NEWS scores were between eight and twelve from Day 33 through Day 45 and then remained below seven from Day 46 until discharge. The patient was weaned off mechanical ventilation on Day 49.

### COVID-19 diagnosis

COVID-19 was diagnosed using the 2019 Novel Coronavirus (2019-nCoV) Real-Time Reverse Transcriptase (RT)–PCR Diagnostic Panel from the International Reagent Resource. A threshold cycle (Ct-value) within 40 is considered COVID-19 positive.

### Virus isolation

SARS-CoV-2 was recovered from each of three nasopharyngeal swab samples, diluted 1:100, and passaged on Vero (NR-10385, BEI Resources) or Vero E6 (CRL1586™, ATCC) cell lines for a maximum of three times until the cytopathic effect was observed.

### Tissue culture infectious dose (TCID_50_)

Viral samples were serially diluted from 1:10^1^ to at most, 1:10^12^ in Opti-Minimal Essential Medium Reduced-Serum Medium. 200µL of the diluted virus was placed in four wells of Vero E6 cells that were seeded in 96-well plates for each dilution for 1 day and incubated at 37°C in 5% CO_2_ for 3 days. Cytopathic effects were recorded. TCID50 represents the viral loads causing a cytopathic effect in 50% of the wells as calculated by the Reed-Muench method^7^.

### Genome sequencing and data analyses

SARS-COV-2 whole genome RT-PCR amplification was conducted by either using Access Array (AA) system (Fluidigm Corporation, CA, USA) with 35 pairs of customer designed specific primers targeting the reference sequence NC_045512.2, or using the two step RT-PCR ARTIC protocol version 3 (24-Mar-2020) with v3 primers pool 1 and pool 2 (https://artic.network/ncov-2019). Amplicon libraries were then prepared using the Nextera DNA Flex Library Prep kit, followed by sequencing with MiSeq Reagent Kit v3 (600-cycle) and MiSeq sequencing system (Illumina, San Diego, CA, USA), as described elsewhere^8^. The quality of paired-end reads obtained from MiSeq sequencing were analyzed by using Qiagen CLC Genomics Workbench 20.0.4 and Qiagen “Analysis of SARS-CoV-2 using MinION sequences” Protocol with a quality threshold of 0.05, and a minimal coverage of 10× was used in genetic variant analyses. Sequence identities were calculated using MEGA v10.1.8 with the function of p-distance, the proportion (p) of nucleotide sites (http://www.megasoftware.net/mega.php). The average coverages of the three samples are 118,392 (Sample 20×1087), 236 (Sample 20×1119), and 4021 (Sample 20×1120) reads. Coverages were 100% (Sample 20×1087), 45% (Sample 20×1119), and 52% (Sample 20×1120) of the reference sequence, NC_045512.2.

### Phylogenetic analyses and molecular characterization

Nucleotide sequences were aligned using MUSCLE v3.8.31, and the mutations were analyzed using BioEdit v7.2.5. The maximum-likelihood (ML) phylogeny tree was constructed using BEAST 2.5 with Yule Model, and Strict Clock. The chain length was 10,000,000 with burn-in rate 10%. The ESS was 902. The posterior probability distributions > 50% are reported in the Figure 1C. Phylogenetic trees were then visualized by Figtree v1.4.4. (http://tree.bio.ed.ac.uk/software/figtree/).

**Figure 1.**
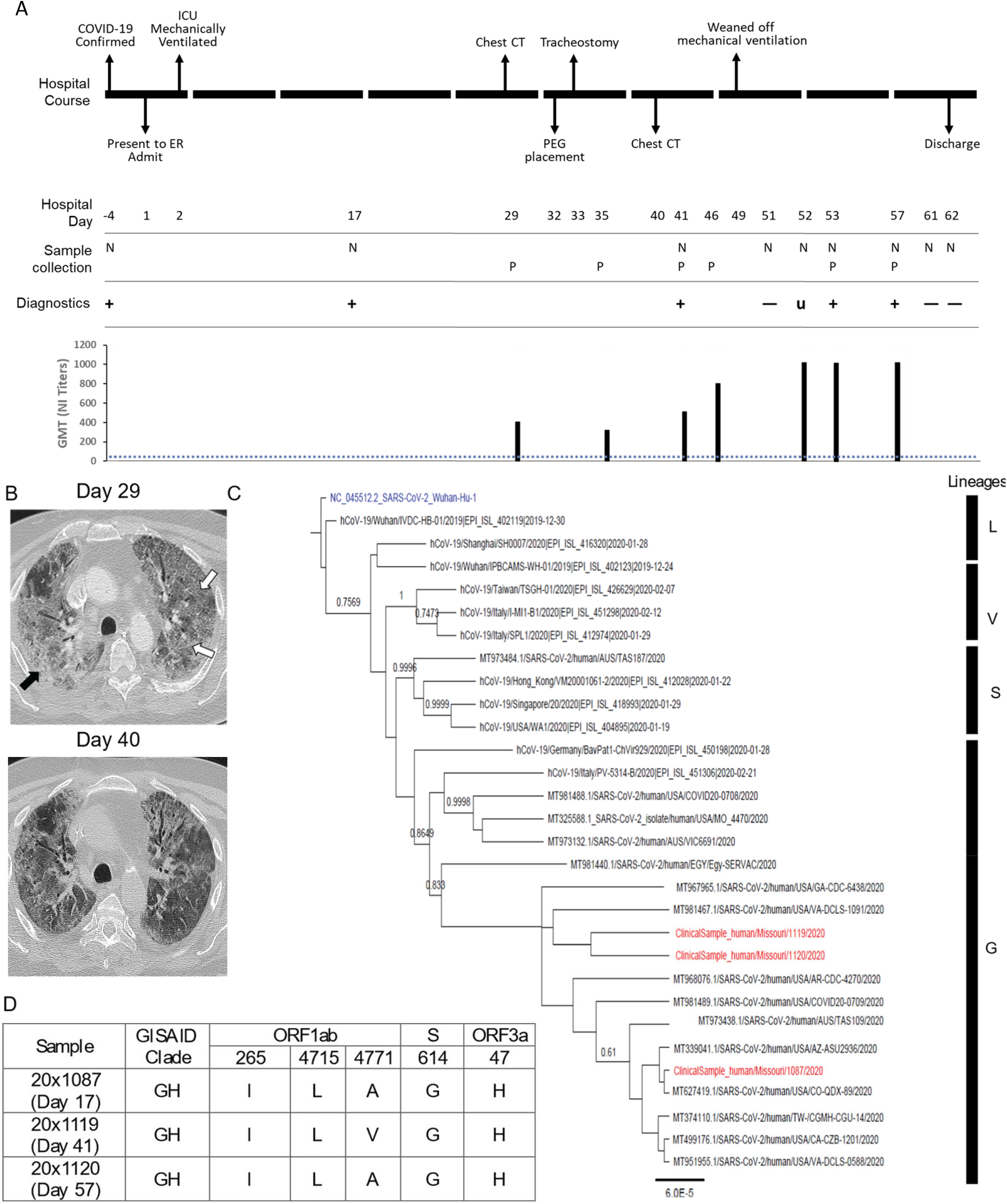
Time course of clinical, virological, and immunological responses in a COVID-19 patient with a prolonged clinical course. A) Clinical outcomes, SARS-CoV-2 infections, and neutralizing antibody responses demonstrate the course of clinical improvement and are associated with the elevation of neutralizing antibodies and occurrence of apparent re-infection; B) CT chest demonstrates diffuse bilateral ground glass opacities (white arrow) and patchy areas of subpleural consolidation (black arrow) (left image, Day 29), and follow-up CT chest (right, Day 40) shows interval improvement in the airspace opacities. ER, emergency room; ICU, intensive care unit; PEG, percutaneous endoscopic gastrostomy; N, nasopharyngeal swab; P, plasma; +, SARS-CoV-2 positive by RT-PCR; -, SARS-CoV-2 negative by RT-PCR; u, SARS-CoV-2 inconclusive by RT-PCR; GMT, geometric mean titer; NI Titers, neutralizing titers. The dashed line denotes the titer of 1:40 which was used to define seroconversion. C) Phylogenetic tree of the viruses with complete genomes. The phylogenetic tree was rooted with the SARS-CoV-2 isolate Wuhan-Hu-1 (Genbank accession No: NC_045512.2) (color in blue). Bayesian posterior probabilities are indicated in the nodes. Scale bar shows the average number of substitutions per nucleotide site. The sequences from three clinical samples are colored in red. D) Summary of amino acid mutations in the viruses from clinical samples 20×1087, 20×1119, and 20×1120.

## RESULTS

### COVID-19 Disease Course

In March 2020, a Caucasian male in his 60s presenting to Urgent Care with fever, weakness, fatigue, rhinorrhea, and cough was diagnosed SARS-CoV-2 positive after three days of disease onset. The patient had recently returned from international travel and had comorbidities of hypertension and hyperlipidemia. Four days later (7 days after disease onset), the patient was admitted to the hospital (Day 1) with increased shortness of breath (Figure 1A). X-ray revealed bilateral, patchy ground glass opacities consistent with viral pneumonia. Hydroxychloroquine and broad-spectrum antibiotics were initiated during the first week of admission.

On Day 2 (D2), the patient developed acute hypoxic respiratory failure and was transferred to the Intensive Care Unit (ICU), requiring endotracheal intubation, mechanical ventilation, and intermittent vasopressors. His hospital course was complicated by secondary bacterial pneumonia, eosinophilic bronchiolitis, and oral candidiasis. A computerized tomography (CT) scan performed on D29 showed extensive diffuse bilateral ground glass opacities and subpleural consolidation consistent with COVID-19-associated respiratory failure (Figure 1B). A percutaneous endoscopic gastrostomy and tracheostomy were placed on D32 and D33, respectively. A follow-up CT chest scan on D40 showed interval improvement in the airspace opacities (Figure 1B).

### Nasopharyngeal swab test results

Viral RNA was undetectable on D51 and inconclusive on D52. A follow-up nasopharyngeal swab was positive again on D53 and a confirmatory nasopharyngeal swab on D57 indicated markedly increased viral RNA load. On D61 and D62, two nasopharyngeal swabs revealed undetectable viral RNA. The patient was discharged on D63 in stable condition.

### Genome sequencing and elevated viral load

Due to the positive-negative-positive results of this patient, the viruses were recovered and sequenced. Viruses were recovered from the clinical samples collected on D17 (Sample 20×1087, threshold cycle [Ct] = 32.5), which was 24 days post disease onset. The sequences recovered for Sample 20×1087 contained 29,904 nucleotides (GenBank accession No: MW004168), Sample 20×1119 contained fragments totaling 13,499 nucleotides, and 20×1120 contained fragments totaling 15,556 nucleotides. The complete genomic sequence of Sample 20×1087 was 99.980% identical to SARS-CoV-2 isolate Wuhan-Hu-1 (Genbank accession No: NC_045512.2). The pairwise nucleotide identities of the three clinical samples were 99.970% (20×1087 vs 20×1119), 99.974% (20×1087 vs 20×1120), and 99.973% (20×1119 vs 20×1120) in the approximately 10 kb overlapping region. Growth assays for these three samples were performed to test for infectious virus, and only the sample at D17 was viable.

The viruses from all three samples belong to the G lineage of SARS-CoV-2 (Figure 1C) with D614G in the S protein, P4715L in non-structural protein (nsp) 12, T265I at nsp2, and Q57H at open reading frame 3a (ORF3a), consistent with the prevalent D614G/Q57H/T265I subclade in the United States ^9^. Limited number of polymorphisms were identified among these three viruses (Figure 1D). Of note, A4771V at ORF1ab was identified only in 20×1119 but not in the other two samples (20×1087 and 20×1120).

### Analysis of neutralizing antibodies

Plasma samples were collected between D29 and D57 for neutralization/inhibition (NI) and ELISA determination (Figure 1A, Table 1). Live SARS-CoV-2-based NI assays showed highly positive and constant Nab titers, ranging from 1:403 to 1:320, until D41. On D52, when the patient retested indeterminate then positive, the Nab geometric mean titer increased to 1:1016, remaining elevated until discharge. Pseudotyped NI results also indicated raised Nab titers on D52.

**Table 1.** Comparison of neutralizing antibodies using microneutralization assays with live SARS-CoV-2 virus, neutralization analyses with pseudovirus expressing spike proteins, and ELISA assays. NI-live assays were performed in triplicate, and geometric mean values were calculated. NI-pseudo assays were performed in triplicate, and titers were calculated using regression analyses to correspond to 50% inhibition. ELISA assays were performed in duplicate, and the mean values were calculated. NI-live, Neutralization inhibition assays using live virus; NI-Pseudo, Neutralization inhibition assays using pseudovirus; -, not available; GMT, geometric mean titer; SD, standard deviation.

| Day | NI-live GMT (±SD) | NI-pseduo (Mean Titers) | ELISA (Mean Titers) |  |  |  |  |  |  |  |  |  |  |  |
| --- | --- | --- | --- | --- | --- | --- | --- | --- | --- | --- | --- | --- | --- | --- |
|  |  |  | RBD |  |  | S (Full length) |  |  | S1 |  |  | S2 |  |  |
|  |  |  | IgG | IgM | IgA | IgG | IgM | IgA | IgG | IgM | IgA | IgG | IgM | IgA |
| 29 | 403 (±1.49) | 8600 | 8100 | 300 | 900 | 8100 | 300 | 900 | 8100 | 100 | 300 | 24300 | 300 | 900 |
| 35 | 320 (±1.00) | 10148 | 8100 | 300 | 900 | 8100 | 300 | 900 | 8100 | 100 | 300 | 24300 | 300 | 600 |
| 41 | 508 (±1.49) | 7287 | 8100 | 300 | 900 | 16200 | 300 | 900 | 8100 | 100 | 300 | 8100 | 300 | 300 |
| 46 | 806 (±1.49) | - | - | - | - | - | - | - | - | - | - | - | - | - |
| 52 | 1016 (±1.49) | 10746 | 8100 | 100 | 900 | 16200 | 300 | 900 | 8100 | 100 | 900 | 8100 | 100 | 300 |
| 53 | 1016 (±1.49) | - | - | - | - | - | - | - | - | - | - | - | - | - |
| 56 | 1016 (±1.49) | - | - | - | - | - | - | - | - | - | - | - | - | - |
| 57 | - | 8773 | 16200 | 100 | 300 | 8100 | 300 | 900 | 8100 | 100 | 900 | 8100 | 100 | 200 |
| Negative control | <1:40 | <1:40 | 50 | 50 | 50 | 50 | 50 | 50 | 50 | 50 | 50 | 50 | 50 | 50 |

ELISA results showed elevated RBD, S, S1, and S2-specific IgM, IgG, and IgA titers for the first 45 days (Table 1). While other antibodies remained stable or began declining following the first negative test, RBD-specific IgG and S1-specific IgA titers continued rising throughout the hospitalization (Table 1). Elevated RBD-specific IgG and S1-specific IgA titers correlated with the patient retesting positive on D52.

## DISCUSSION

In current CDC interim guidance on duration of isolation and precautions for adults with COVID-19, persons with severe or critical illness are recommended to be removed from transmission-based precautions 20 days after initial disease onset, at least 24 hours after their last fever without fever-reducing medications, and improved symptoms^6^. In this case report, we show that it is possible for a not severely immunocompromised patient with severe and persistent COVID-19 symptoms to continue shedding infectious virus beyond 20 days after symptom onset, even after being afebrile for multiple weeks. This patient presented with viable virus growth, detected in the laboratory, 24 days after symptom onset, suggesting that patients with severe and persistent COVID-19 may have a longer viral infectivity than originally realized.

The patient also presented with fluctuating viral loads and increased antibody titers during severe acute infection and extended persistent infection. The boosted Nab titers and viral RNA from nasopharyngeal swabs on D52 after a two-day RNA-negative result suggests the patient may have experienced a recurrent infection during his prolonged hospitalization. Viral shedding in COVID-19 patients can last up to three months after disease onset^10^. With high sequence identities among three viruses and a limited number of polymorphisms (Figure 1D), the second infection was likely a recurrent infection. It is possible that the second infection could be caused by reactivated viruses in this patient, although the patient’s symptoms improved.

The primary limitation of this study is that the available data encompasses a single patient. Confounding factors include the multiple clinical treatments that the patient received. Bacterial pneumonia was treated with broad spectrum antibiotics from D3-40. Antifungals were given to treat oral candidiasis D45-52, and methylprednisolone (steroid) was administered from D41-57 for eosinophilic bronchiolitis. In summary, the patient was on steroid medication and had just completed antifungal therapy when they retested positive for SARS-CoV-2 between D54-56. Additionally, previous studies indicate T-cell immune responses, which were not analyzed in this study, might play roles in clinical outcomes.

A strength of this study is the use of three serological assays to monitor antibody development. Intriguingly, only S1-based IgA titers clearly increased via ELISA during the recurrent infection period, consistent with Nab titers using live virus (Table 1). Although the pseudotyped NI assay detected Nabs, potential discrepancies from live virus-based NI assay, as in this study, indicate that caution is needed when interpreting pseudotyped NI data.

In summary, this report follows the clinical and serological timeline of a patient with severe and persistent COVID-19. This patient demonstrated viable virus growth at least 24 days after symptom onset. These findings suggest that separate discontinuation of transmission-based precaution guidelines for patients with persistent symptoms and extended hospital stays may be necessary.

## Data Availability

Data available on request from the authors

## Acknowledgments

We are grateful for support from Jeffery Adamovicz, Travis McCarthy, Paul Anderson, and Jeff Whyte at University of Missouri-Columbia.

## Author contributions

Manuscript draft: X.F. W., C. Y. T., and D. R. ; Manuscript revision: H. X., J. H., J. A. M., N. C., and R. H.; Data analyses: C. Y. T., X. F. W., and D. R.; Virological analyses: Y. W.; Serology: K. S., M. J. M. K., H. J. K., and H. X.; Genomic sequencing: T. L., J. H., and N. D. C.; Pathology: D. R., R. D. H., and S. C.; Radiology: A. H.

